# Wastewater based surveillance system to detect SARS-CoV-2 genetic material for countries with on-site sanitation facilities: an experience from Bangladesh

**DOI:** 10.1101/2021.07.30.21261347

**Authors:** Md. Jakariya, Firoz Ahmed, Md. Aminul Islam, Tanvir Ahmed, Abdullah Al Marzan, Maqsud Hossain, Hasan Mahmud Reza, Prosun Bhattacharya, Ahmed Hossain, Turasa Nahla, Newaz Mohammed Bahadur, Mohammad Nayeem Hasan, Md. Tahmidul Islam, Md. Foysal Hossen, Md. Didar-ul-Alam, Nowrin Mow, Hasin Jahan

## Abstract

The presence of SARS-CoV-2 genetic materials in wastewater has become a matter of grave for many countries of the world. Wastewater based epidemiology, in this context, emerged as an important tool in developed countries where proper sewage system is available. Due to the recent shift in the spread of the infection from urban to rural areas, it is now equally important to develop a similar mechanism for rural areas as well. Considering the urgency of the issue a study was conducted in 14 districts of Bangladesh and a total of 238 sewage samples were collected in two different periods from December 2020 to January 2021. We are the first to propose a surveillance system for both urban and rural areas where a proper sewage system is absent. Based on RT-PCR analysis of the water samples, in more than 92% of cases, we found the presence of the SARS-COV-2 gene (ORF1ab, N, and Internal Control-IC). The trend of Ct value varies for different study locations. The spread of genetic material for on-site (Δm = 0.0749) sanitation system was found more prominent than that of off-site sewage system (Δm = 0.0219); which indicated the shift of genetic material from urban to rural areas. Wastewater samples were also measured for physicochemical parameters, including pH (6.30 - 12.50) and temperature (22.10 - 32.60) ºC. The highest viral titer of 1975 copy/mL in sewage sample was observed in a sample collected from the isolation ward of the SARS-COV-2 hospital. Additionally, a correlation was found between bacterial load and SARS-CoV-2 genetic materials. The results indicated the association of increased Ct values with decreasing number of patients and vice versa. The findings reported in this paper contributed to the field of wastewater-based epidemiology dealing with SARS-COV-2 surveillance for developing countries where proper sewage system is absent and highlighting some of the challenges associated with this approach in such settings.

**Highlights:**

- Development of wastewater-based surveillance system based on on-site sanitation system for developing countries.
- Association of different environmental parameters with the presence of SARS CoV-2 genetic material in wastewater.
- Prediction of the viral concentration of sewage system using viral load and copy number parameter.

**Graphical Abstract:** 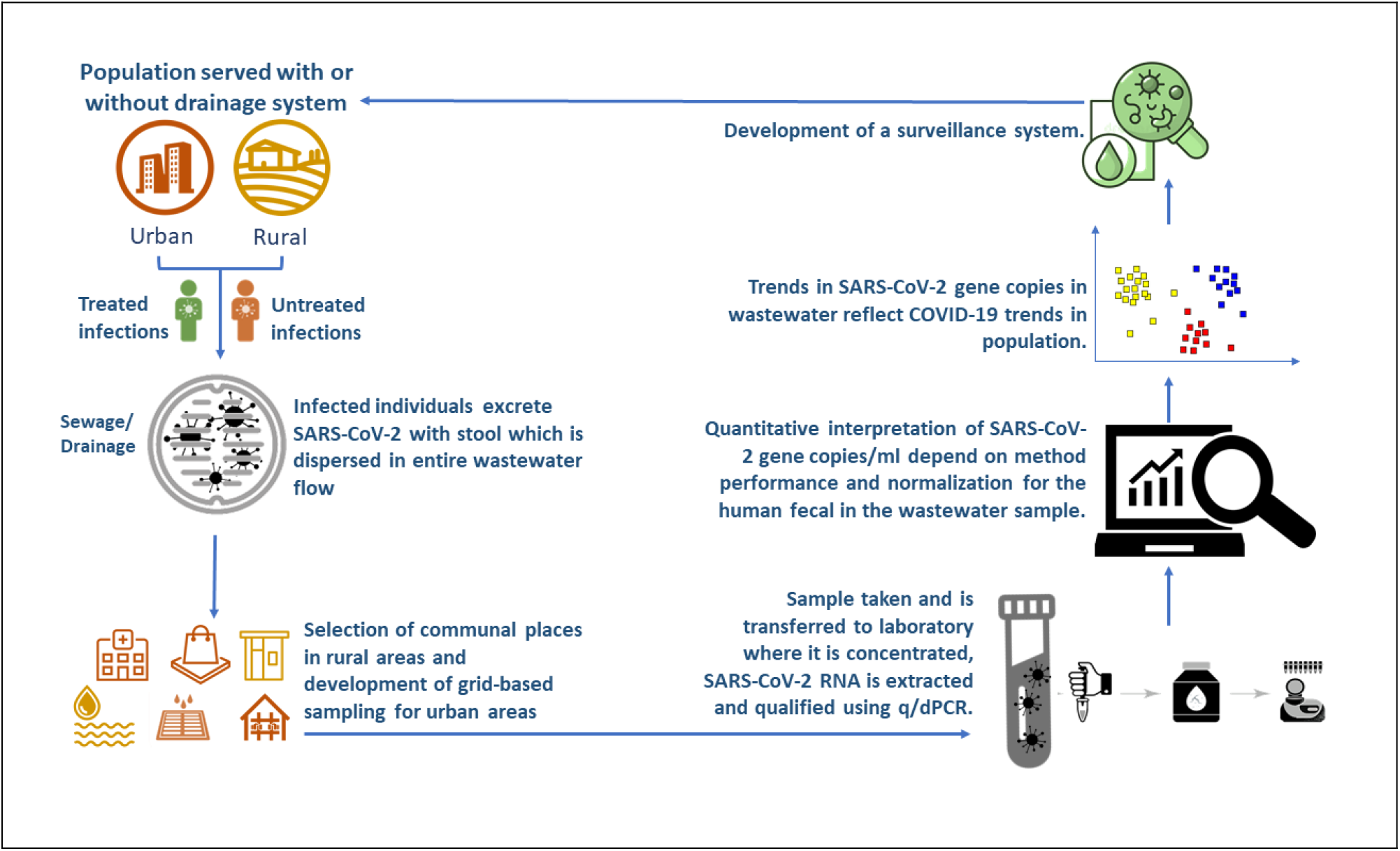

## 1. Introduction

The novel coronavirus (SARS-CoV-2) has evolved as a pandemic by rapidly spreading worldwide (Mao, et al., 2020). The dynamic nature of the SARS-CoV-2 along with massive confirmed cases each day has made it difficult for the world to detect through individual tests (Ott, 2020). In this particular context, wastewater based epidemiology (WBE) emerged as a significant tool in many countries to detect SARS-CoV-2 genetic material in the wastewater (Ammar et al., 2021; W. Ahmed, Angel, et al., 2020; Haramoto et al., 2020; La Rosa et al., 2020; Lodder & de Roda Husman, 2020; Medema et al., 2020; Nemudryi et al., 2020; W. Ahmed, Angel, et al., 2020). However, the WBE was particularly efficient for the countries where a proper sewage system is available (off-site sanitation system). Globally, there are around 45% of the population who have access to a proper sewage system, which can prevail over the wastewater surveillance system (World Health Organization, 2019). On the contrary, 90% of the sanitation facilities in the developing countries which are primarily on-site based, cannot be benefitted from the WBE system (Sagoe, et al., 2019; Rahman, et al., 2016; Diaz & Barkdoll, 2012).

Bangladesh is a developing country that is a fusion of both off-site and on-site sanitation systems. Dhaka, the capital city, only consists of an off-site sanitation system to which only 20% population is connected (Masour, Islam, & Aktaruzzaman, 2017; Rahman, et al., 2016) and other areas in Bangladesh depends on the on-site sanitation system. This versatility of the country makes it difficult to primarily depend on the methods of the WBE surveillance system which is efficient for a centralized sewage system. It is also important for developing countries like India, Pakistan, Africa, and other countries with similar sanitation facilities, to establish a WBE surveillance system to monitor the prevalence of the disease.

Recently, the nature of the pandemic has evolved in both the urban and rural areas which rely on on-site sanitation facilities (Ekumah, et al., 2020; Wang, et al., 2021). In these areas, the current boost in the statistics of SARS-COV-2 confirmed cases have been mostly identified through individual tests which are based on huge expense and time whereas similar identification is possible by developing a wastewater surveillance system. A methodology to detect SARS-Cov-2 through WBE is published previously in numerous papers, targeting the first world countries with the proper infrastructure of the sewage facilities. (Orive et al., 2020; Prevost et al., 2015; Randazzo et al., 2020; Rimoldi et al., 2020; Sherchan et al., 2020; Wu et al., 2020; Wurtzer et al., 2020; Tang et al., 2020).

We are the first to propose a surveillance system for both urban and rural areas where a proper sewage system is absent and intending to contribute to policy decisions regarding the WBE inclusion in developing countries by identifying sampling methods for both urban and rural areas. Which is primarily based on an earlier method which we developed (Ahmed et al., 2021). The present study is carried out to develop a proof of concept to develop and roll-out a system of national surveillance of the prevalence of SARS-CoV-2 infections and involves preliminary testing, optimization, and validation of sampling and virus analysis methods, as well as results interpretation and reporting in the context of Bangladesh. This paper is focusing on developing a model to predict the presence of the SARS-COV-2 genetic material in wastewater in countries based on on-site sanitation facilities for both urban and rural areas. The results are likely to appeal to the policymakers worldwide especially of the developing/low sanitation countries depending primarily on on-site facilities to adopt the wastewater surveillance for the SARS-COV-2 pandemic

## 2. Material and Methods

### 2.1. Selection of Sampling Sites

It was immensely difficult for a wastewater surveillance system to function properly in areas with an on-site sanitation facility in the rural areas compared to an off-site sewage system in the urban areas. Hence, the study developed separate methods to collect samples for both on-site and off-site sanitation systems. In settlements that consist of a sewage system (off-site sanitation system), the samples were collected from the drains of Isolation Centers, Hospitals, Prisons, Community Drain and City Drain. (Fig. 1). On the other hand, in areas lacking sewage system (on-site sanitation system), the sampling sites were selected by identifying the wastewater channels around the nodal points like mosques, railway stations, bus stations, communal ponds, and other public locations as hotspots to represent the whole community.

**Fig. 1.**
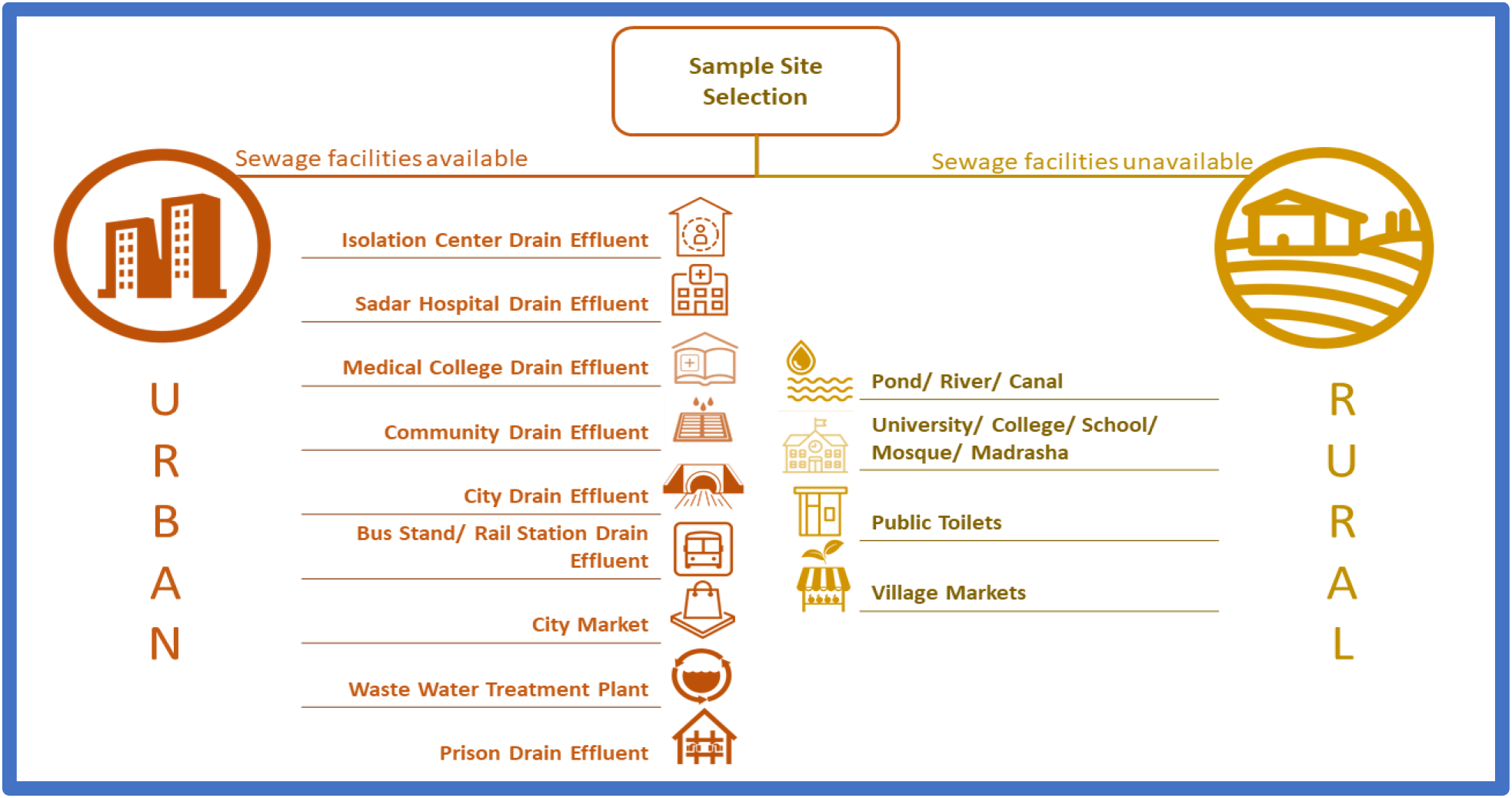
Specific sampling points were selected for wastewater-based epidemiology based on sewage facility. Drain effluent, city market, treatment plants were selected where proper sewage system is available (off-site sanitation system). Whereas pond, river, public toilets, village markets were selected where proper sewage system is unavailable (on-site sanitation system).

#### Grid-Based Sampling

For urban areas where a sewage system is present, we first located the whole drainage system of the area. Later, with the help of GIS software, we separated the households into grids to understand the cluster of houses and conclude the best sampling points from the drainage. (Supplementary Fig. 1).

#### Hotspot-Based Sampling

In terms of rural areas, the sampling method was different as there were no sewage facilities available. Thus, we followed a hotspot-based sampling where we selected some nodal points which are mostly used by rural publics such as mosques, rail stations, bus stations, communal ponds, and canals.

### 2.2. Sampling

A total of 238 wastewater samples were collected from 7 divisions, 14 districts that represented the off-site and the on-site facilities of Bangladesh (Fig. 3). We collected repeated samples from the same place in 15 days duration while the first round was completed from 1^st^ December to 15^th^ December 2020 and the second round completed 1^st^ January to 15^th^ January 2021(Supplementary Table-1). This is to be mention-worthy that wastewater specimens were collected from various locations of the capital Dhaka City such as Kuwait Moitri Hospital, Mugda Medical College and Hospital, and Kurmitola General Hospital, Mohammadpur, Korail Slum, and Mirpur Slum. Specific locations were used for sample collection that continued two times during the tenure from November 2020 to January 2021. Samples were collected in 50ml sterile falcon tubes and carried in the icebox. All wastewater specimens were refrigerated at 4^°^C and brought to the laboratory for further analysis. Sterile falcon tubes were used for sampling, and blanks in the same group were analyzed to determine if there is any contamination during the transport. During sampling time, various external factors were counted. All the experiments and analyses were carried out at the SARS-COV-2 Diagnostic Lab, Dept. of Microbiology, Noakhali Science and Technology University.

As it has been determined in several studies (Chen et al., 2021; Kaplin et al., 2021) that environmental factors play a significant role in SARS-COV-2 pandemics, the pH and temperature of samples were measured with a portable pH meter (Milwaukee) and thermometer (TP-300) respectively during the collection of samples.

For total bacterial count, the nutrient agar plating method was performed following microbiological standard protocol with serial dilutions of the wastewater in normal saline to predict the most probable number of inhabiting bacteria.

### 2.3. Sample Preparation, Concentration, and RNA Extraction Procedure

First of all, samples were filtered using Whatman filter paper to remove large particles according to the same concentration procedure, as described by Kumar et al (Kumar et al., 2020). Briefly, sewage samples (50 mL) were centrifuged (Thermo Scientific) at 4500×g for 30 min followed by filtration of supernatant using 0.22-micron filters (Himedia). Further, each sewage filtrate was concentrated using the polyethylene glycol (PEG) method. In this method, PEG 6000 (80 g/L) and NaCl (17.5 g/L) were mixed in 25 ml filtrate, which was then incubated at 17°C in 100 rpm shaking overnight. The next day, the mixture was centrifuged at 13000×g for 90 min. The supernatant was discarded after centrifugation, and the pellet was suspended in 300 μL RNase-free water. This was further used as a sample for viral RNA extraction using a commercially available QIAamp Viral RNA Mini Kit and FavorGen viral RNA extraction kit was used for confirmation of previous extracted RNA. Besides, all the experiments were performed three times for confirmation of the results and accepted where variations were less than 10%. SARS-COV-2 positive patient samples were used as a positive control case in each run. We initially employed qualitative measurement, and hence, increasing and decreasing viral load was measured based on the Ct value. RNA concentrations were measured by NanoDrop (Thermo Scientific TM NanoDrop 2000 and 2000c, BioRad) and were stored at -70 °C until further use.

### 2.4. RT-PCR Analysis

To detect SARS-COV-2 confirmed positive patients, the Bangladesh Directorate General of Health Service (DGHS) expert panel selected two genes (ORF1ab, N) based SARS-CoV-2 virus detection system with human RNAse P gene as the internal control (IC) of the RT-PCT system. Hence, the same genes were used for SARS-COV-2 genetic materials detection from sewage samples.

RNAs were analyzed for the detection of SARS-CoV-2 by RT-PCR (CFX96, BioRad) using the RT-PCR kit (Sansure Biotech Inc., China). As described in the product manual, technical procedures were carried out, and interpretations of results were made. In brief, we had set the sample layout with RT-PCR protocol covering 45 cycles containing FAM fluorescence select for ORF1ab, ROX for N gene as well as CY5 for human RNase P gene as the internal control (IC). Ct value (cycle of threshold) is assigned as quantitative value and used for detection of various gene expressions, copy number, and viral load. As quality control measures, positive and negative control were also included in each run to validate the test procedure. For RT-PCR result validation we used another BGI RT-PCR kit. The Novel Coronavirus (2019-nCoV) Nucleic Acid Diagnostic Kit (PCR-Fluorescence Probing) is a real-time reverse transcription-polymerase chain reaction (RT-PCR) test. The 2019-nCoV primer and probe set(s) were designed to detect RNA from SARS-CoV-2.

All test controls (Positive and Negative control) were verified before the interpretation of results. If the controls were not valid, the results were not interpreted as per standard experimental protocol. Instructions for cutoff value and curve shapes for positive, weakly positive, and negative samples after amplification were strictly reviewed.

### 2.5. Copy Number and Viral Load (virus concentration)

Viral gene copy was calculated according to this formula given below:

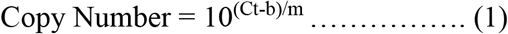

where, Ct, b and m represents the cycles of the threshold value, constant of log 10 based straight line (derived from the regression plot of Ct value associated with the standard copy number inputs) and the tangent of the same straight line. Whereas the viral load (viral gene copies/L of wastewater) was determined according to this formula:

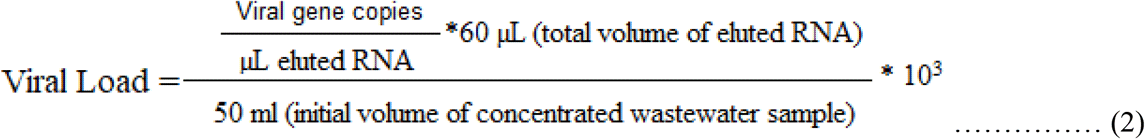

### 2.6. Result Validation

For RT-PCR result validation another RT-PCR kit was used randomly to verify the results of both positive and negative samples obtained by the regularly used kit (Supplementary Fig. 2). Similarly, the RNA extraction process was assessed by another viral RNA extraction kit where all previous results were found comparable. We also compared our results with the positive national cases and found similarities with only a 3-5% deviation. Therefore, considering two crucial findings from our pilot initiative: identifying sampling location and the test results, we can confidently propose the method of developing rural wastewater-based epidemiology (RWBE) for rural areas of the developing world.

## 3. Results

### 3.1. Comparison of First and Second Round Sampling

The distribution of SARS-CoV-2 genetic materials in all sampling regions indicates the presence of confirmed SARS-COV-2 cases in entire sampling areas. In both (1^st^ time and 2^nd^ time) sampling rounds, the lowest existence of IC gene and the highest occurrence of ORF1ab gene were found. A total of 46 samples were positive for IC gene from first-round sampling whereas 55 samples were positive during the second round. The number of positive samples for N and ORF1ab genes were (62,63) and (56,63) respectively (Fig. 2a). Among the total 238 sewage samples, the distribution of ORF1ab, N, and IC genes were 36%, 35%, and 29% respectively (Fig. 2b)

**Fig. 2.**
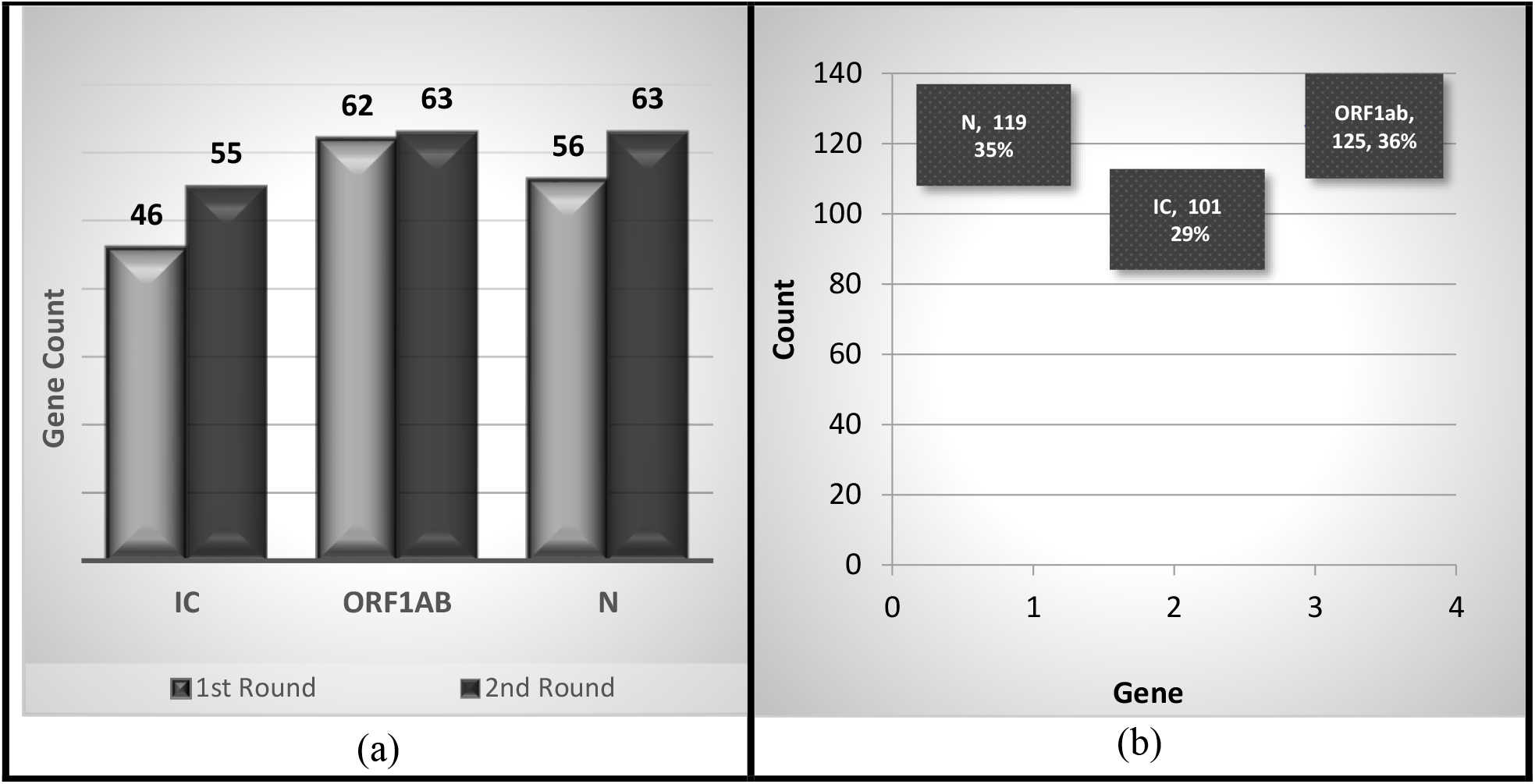
Comparative view of gene pattern (IC, ORF1ab, N) found in wastewater during 1^st^ and 2^nd^ round sampling. **(a)** Comparison (gene count) of target genes out of 238 samples based on 1^st^ and 2^nd^ round result. **(b)** Comparison (percentage) of target genes out of 238 samples (1^st^ and 2^nd^ round combined result).

From the first-round sampling, mean Ct values for the three genes were 35.97 (IC), 35.92 (ORF1ab) and 35.74 (N) with corresponding standard deviations (SD) found 2.78 (IC), 2.55(ORF1ab), and 2.54 (N) compared to second round mean values 36.83 (IC), 36.32 (ORF1ab), 36.04 (N) with SD observed 1.66 (N), 1.93 (ORF1ab), 2.12 (N) respectively. These results indicated that the genetic materials of the 2^nd^ round sewer samples were higher than the 1^st^ round specimens.

### 3.2. Spatio-Temporal Distribution of the SARS-CoV-2 Genetic Materials

During the first-round sampling, a total of 119 wastewater samples were collected from 14 districts of 7 divisions and 114 samples (95%) were found positive for various genes. All three genes (ORF1ab, N, and Internal Control-IC) were found in 26 samples (21%); N gene and IC in 15 samples (12%); ORF1ab and IC in 22 samples (18%). The rest of the samples showed positive for one of the three target genes (Fig. 3).

**Fig. 3.**
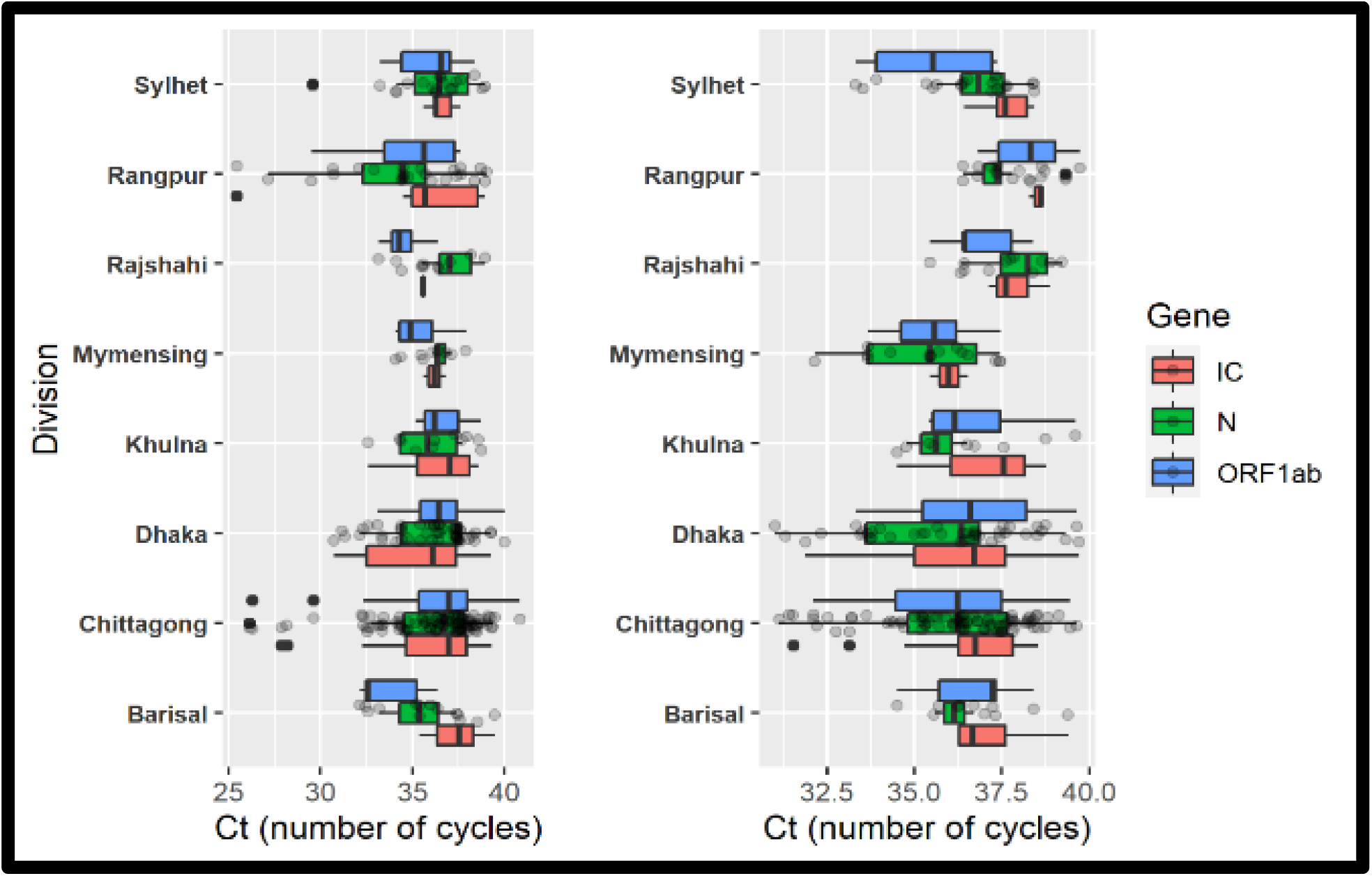
Distribution of ct values in seven divisions. The left portion of the graph represents the first-time sampling (1^st^ December to 15^th^ December 2020) whereas the right portion of the graph represents the second-time sampling (1^st^ January to 15^th^ January 2021). Chittagong, Rangpur divisions had lowest Ct value range in 1^st^ round sampling whereas Dhaka, Chittagong, Mymensing had the lowest in the 2^nd^ round sampling

After analysis of the second-round samples, 117 specimens were found to be positive (98%). The three genes; ORF1ab, N, IC were found in 12 samples (10%), with N and IC genes in 23 samples (19%), and ORF1ab, IC genes in 23 samples (19%) among the total 119 wastewater samples. Therefore, during both rounds, the presence of SARS-COV-2 genetic material was identified in urban areas as well as in rural areas.

Dhaka city showed the highest number of SARS-CoV-2 positive genes in wastewater samples collected both in the first and second round sampling from 14 districts representing 7 divisions of the country. Among different genes, the presence of IC gene (11, 10), ORF1ab gene (11, 10), and N gene (16, 12) were identified during the first-round and second-round sampling respectively. From the first-round sample collection, the highest Ct value for the ORF1ab gene was found at Rohingya Camp (39.37) (both on-site and off-site sanitation system), N gene at Kishorgonj Isolation Center Drain Effluent (40), IC gene at Barisal Isolation Center Drain Effluent (39.48). In contrast, the lowest ORF1ab, and N genes were correspondingly found at Chittagong Isolation Center Drain Effluent (26.14; 26.28), and IC gene at Gaibandha Isolation Center Drain Effluent (25.45) (as a part of Off-site sanitation system). Hence, according to the demographic illustration predominant genetic materials were observed at Chittagong and Gaibandha districts compared to others during the first-round sampling. In the samples, collected during the second round, the highest Ct value was observed for ORF1ab gene at Rangpur Sadar Hospital Drain Effluent (39.73), and N gene at Rohingya Camp (39.65), with IC gene at Kurmitola General Hospital Drain Effluent in Dhaka (39.7) while the lowest ORF1ab and N genes found at Rohingya Camp (32.11), and N gene at Kurmitola General Hospital Drain Effluent (31.00) in Dhaka, with IC gene at Rohingya Camp (31.53). It was also observed that the Rohingya Camp samples showed the highest prevalence of SARS-CoV-2 genes in the samples collected during the second round.

### 3.3. Comparative View of Wastewater Samples in Urban (on-site and off-site sewage system) and Rural Areas (on-site sewage system)

As the capital, Dhaka is known as the center of all kinds of business where people come from different parts of the country and world. Wastewater samples were collected from different locations with diverse socioeconomic backgrounds-Korail slum (on-site), Mohammadpur (on-site), Mirpur (on-site), Gulshan (off-site). Interestingly, it was found to have the highest number of genetic materials of SARS-COV-2 compared to other regions in Bangladesh. The Ct value ranges in the first and second round sampling were comparable, as to be mentioned IC gene (30.72-38.44), ORF1ab gene (32.12-37.61), N gene (33.11-37.48) in the first round while IC gene (31.8-39.7), ORF1ab (33.3-38.75), N gene (31.0-39.32) in the second-round samples

In both the first and second round sampling, 13 sewage samples from Isolation centers, 13 samples from Sadar hospital, and 13 from medical colleges were collected for analysis. The most important result from the experiment was the Ct values of the wastewater samples from isolation centers’ sewage samples that trailed the main source of SARS-COV-2 genetic materials as the highest genetic material concentrations among all results obtained in the present study. We found the lowest Ct IC (25.45), ORF1ab, and N gene (26.14 and 26.28) from Gaibandha (on-site) and Chittagong (on-site) isolation centers respectively.

The viral load of the 2^nd^ round samples showed a negative trend compared to the 1^st^ round sampling at on-site sanitation system (1^st^ round, m= 0.0073; 2^nd^ round m= -0.0822) (Fig. 4a). On the other hand, at off-site sewage systems, the presence of genetic material was also found higher in the 2^nd^ round sampling (m= -0.0283) than that of 1^st^ round sampling (m= 0.0064) (Fig. 4b) However, the increasing trend of viral load at the off-site sewage system was less predominant than that of the on-site sanitation system.

**Fig. 4.**
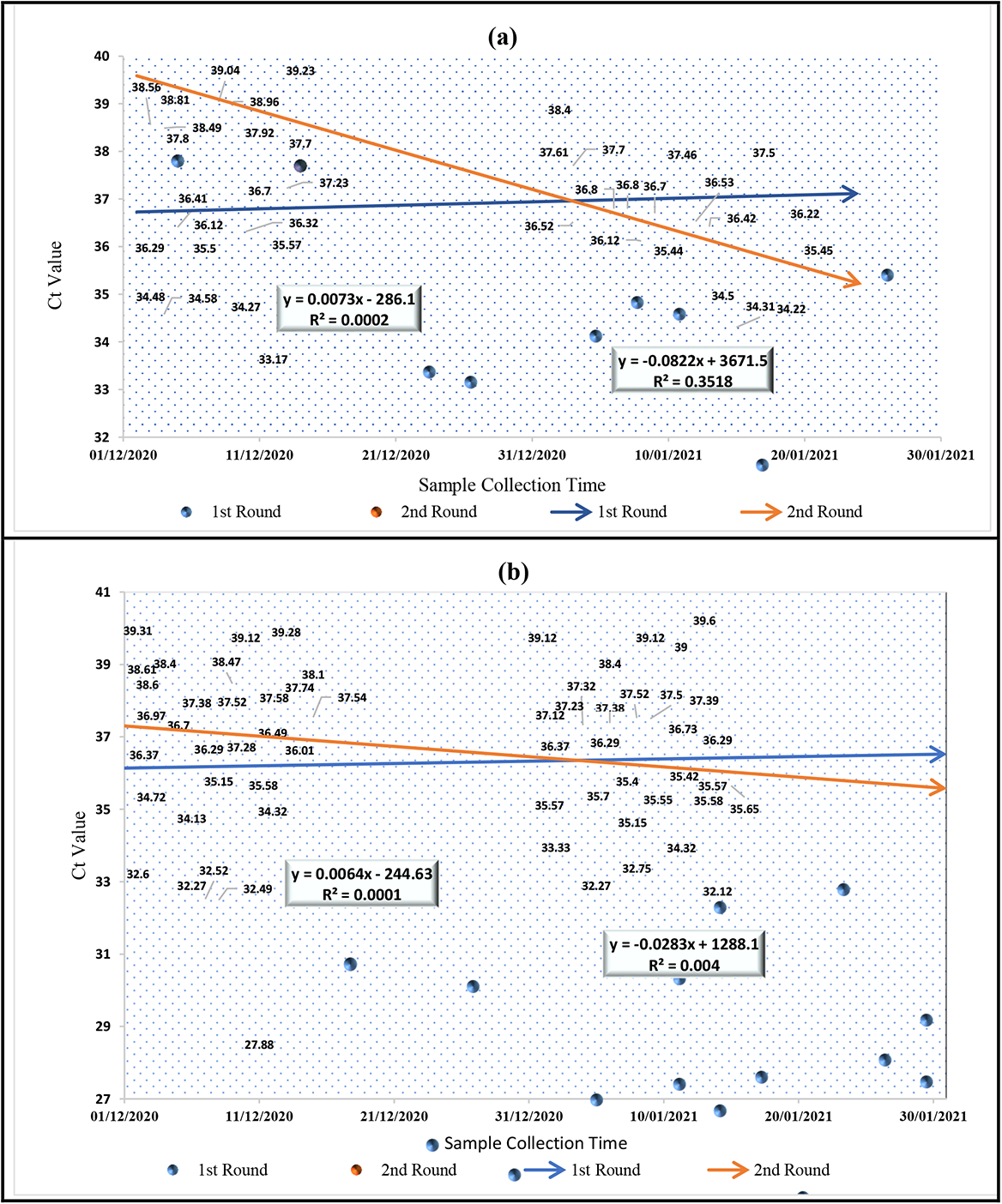
Comparison of Ct value of IC, ORF1ab and N gene found in wastewater in urban and rural areas during 1^st^ time and 2^nd^ time of sampling. **(a)**Comparative lower Ct value during 2^nd^ round of sampling collected from on-site sewage system. **(b)** Comparative lower Ct value during 2^nd^ round of sampling collected from off-site sewage system.

When the first and second round viral load were compared, the spread of genetic material in the on-site (Δm = 0.0749) sanitation system was found more prominent than that of the off-site sewage system (Δm = 0.0219). This trend indicated the shift of genetic material from urban to rural areas

### 3.4. Environmental Factors: Effect of Temperature and pH on Viral Ct value

When considering the effects of temperature and pH on SARS-COV-2 genetic materials distribution in wastewater, strong correlations were found between Ct values and the environmental factors. An increase in temperature was associated with a lowering trend of Ct values, i.e., the upsurge of the genetic material concentrations. Most of the SARS-CoV-2 genes were found at 27^°^C -29^°^C. Similarly, a higher presence of genetic materials was also identified in the acidic to neutral pH (6.8-7.0) ranges compared to alkaline pH. As observed, pH 6.2-7.3 and temperature 23^°^C -32^°^C were the optimum ranges for the maximum presence of SARS-COV-2 genetic materials indicating an interplay between temperature and pH, which has a significant impact on SARS-CoV-2 genetic material concentrations in various sewer samples.

The correlation coefficient indicates that for every additional unit in temperature during first-round sampling, pH, Cy5-IC, FAM-ORFlab, and ROX-N increased significantly by 0.221, 0.367, 0.746, and 0.379, respectively. In addition, FAM-ORFlab is also positively correlated (0.272) with pH and ROX-N gene. Except for the temperature, Cy5-IC showed no relation with other variables.

### 3.5. Association of SARS-COV-2 genes with confirmed cases

According to the study results, Ct values for SARS-CoV-2 genetic material found in the second-round wastewater samples were higher than that of the first-round sampling of wastewater. The study findings showed an increasing trend of SARS-CoV-2 genetic materials in wastewater which was confirmed by the increasing prevalence of SARS-COV-2 positive cases during 2^nd^ round of sampling (January 1^st^ to January 15^th^)

Based on the obtained results from different regions of Bangladesh, it can be concluded that viral Ct values were found highly correlated with the number of cases (Fig. 5)

**Fig. 5.**
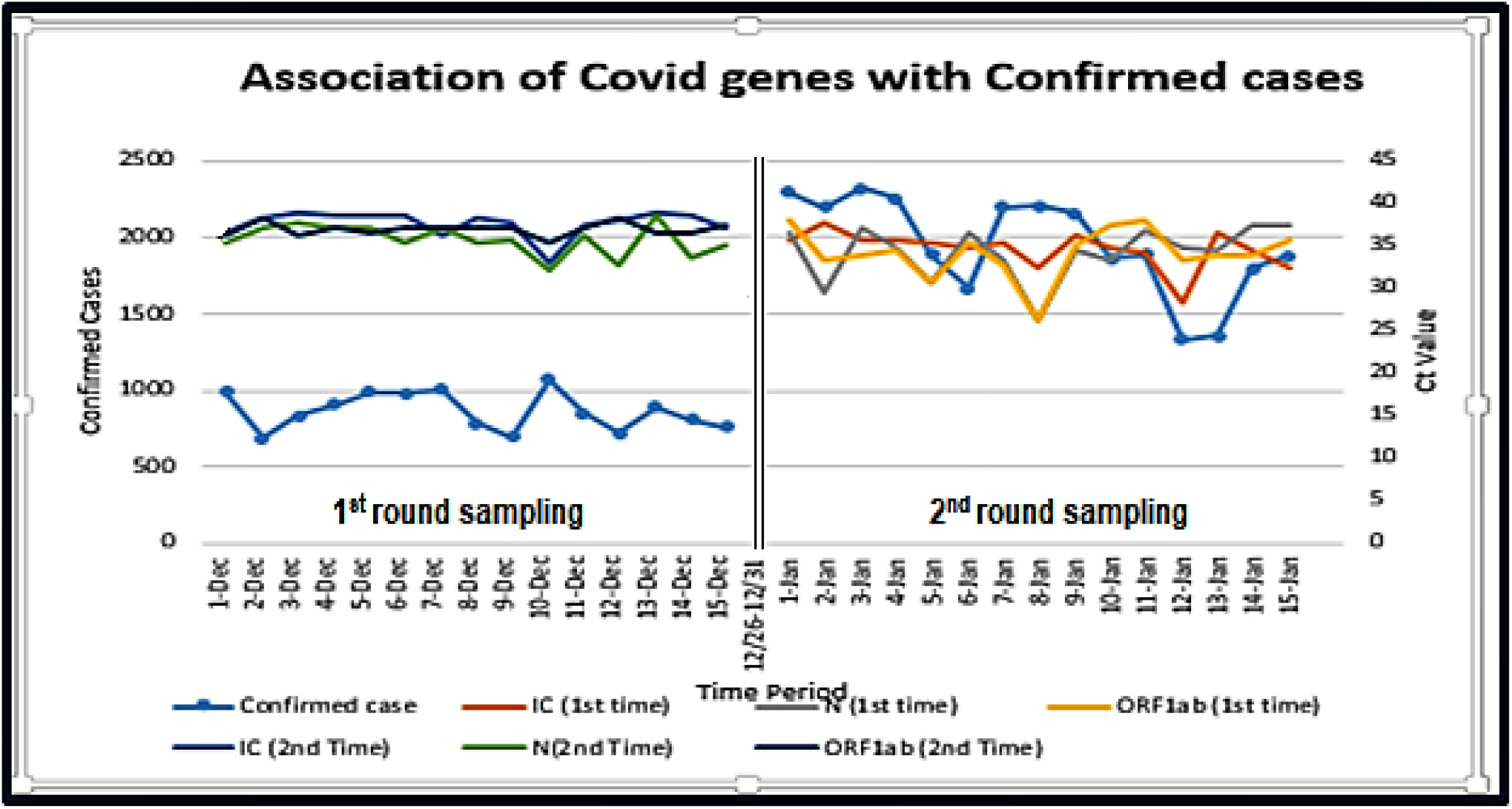
Association of Ct value of three genes (IC, ORF1ab, N) with confirmed cases in two different phases (during 1^st^ time and 2^nd^ round of sampling). As the Ct value increased in the 1^st^ phase the confirmed cases decreased whereas in the 2^nd^ phase the confirmed cases increased as the Ct value decreased.

### 3.6. Total Bacterial Count in Wastewater Samples: Correlation of Bacterial Load with SARS-CoV-2 Genetic Materials

It was identified that the bacterial numbers have a correlation with SARS-COV-2 Ct-value as a higher total bacterial count was found in Dhaka city drains compared to lower numbers at the Rohingya Camp. It is noteworthy that Dhaka city specimens represented the highest number of SARS-CoV-2 genes (100%) with the lowest presence observed in the Rohingya Camp sewer specimens as collected from the community and the wastewater treatment plant. The range of bacterial count in this study was 2.00E+06 to 1.40E+04. More specifically, the total bacterial count was found to have a strong correlation with viral Ct value. It was observed that all genes expressed in the limit of 4.00E+10 to 1.00E+11 bacterial counts.

### 3.7. Viral Concentration (Copy Number and Viral Load)

The result interpretation reminds a strong correlation of viral concentration with the Ct value and hence it is identified as an important factor for viral propagation. From different studies, it has been confirmed that the copy number and the viral load are inversely proportional with the Ct value of the experimented samples. In our study, the copy number fluctuates from 1.00E2 to 1.00E5 whereas, the viral load fluctuates from 1.00E3 to 1.00E7 (Fig. 6). The more the viral concentration increases, the greater the chances of being positive with SARS-CoV-2.

**Fig. 6.**
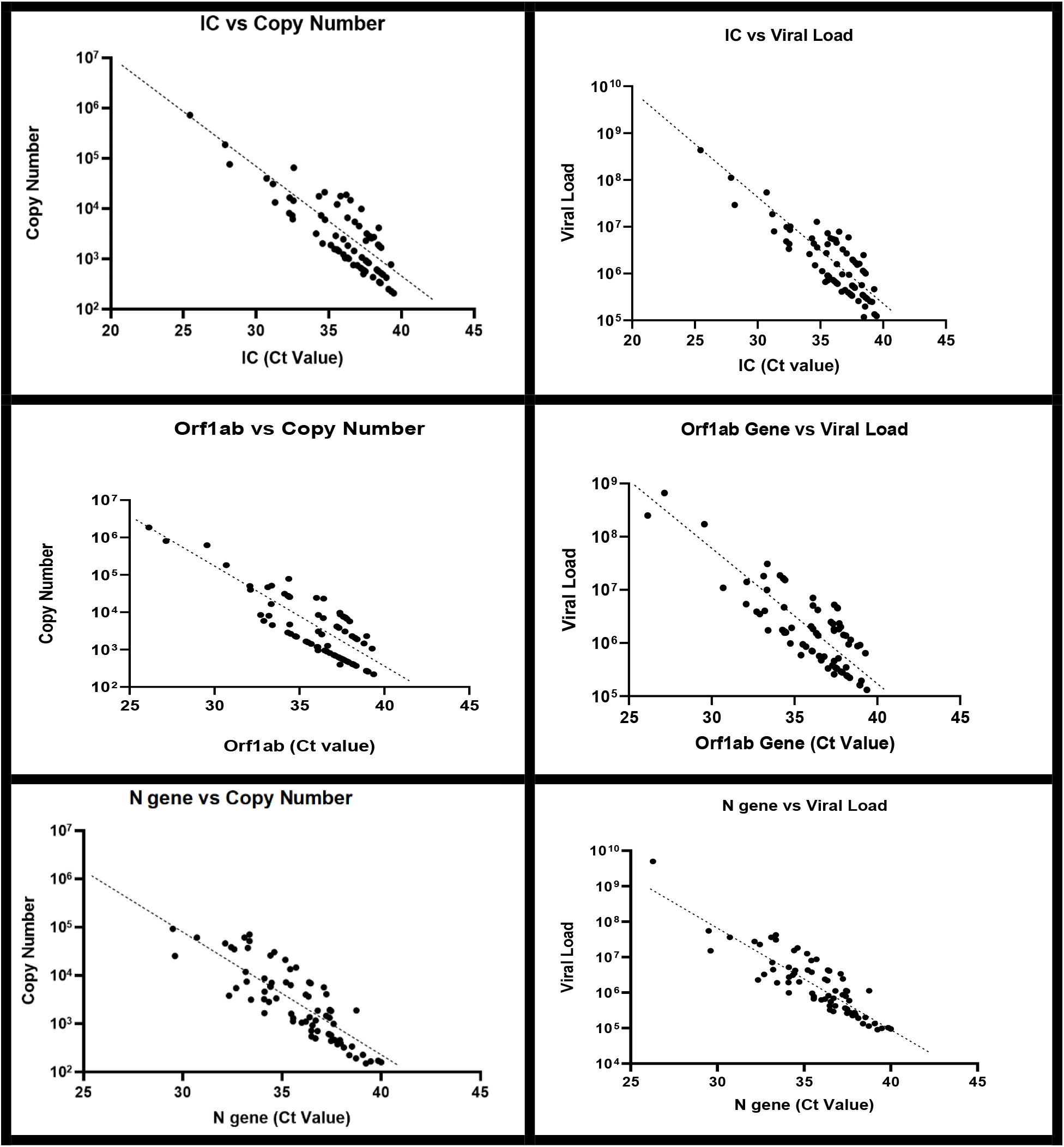
Comparative view of viral load and copy number of three genes (IC, OEF1ab, and N) in on-site and off-site sanitation system. According to equation (1) & (2) viral load and copy number are directly proportional to Ct value but in practical viral load and copy number fluctuate a little bit.

## 4. Discussion

Wastewater surveillance of SARS-CoV-2 provides a powerful tool to evaluate disease incidence at the community level where a proper sewage system is available (Ahmed, et al., 2020; Arora et al., 2020; Prado et al., 2021). However, considering the present spread of SARS-COV-2 from urban to rural areas, it has become important to develop an equivalent system for rural areas as well. Rural wastewater-based epidemiology (RWBE) could be an alternative cost-effective surveillance tool for the management of the SARS-COV-2 pandemic in developing countries. No studies used this methodology to analyze the SARS-CoV-2 spread and the CFR (Case Fatality Rate) of most of the portion of the unadvanced world (where sewage system is unavailable and not well planned). It is therefore clearly demonstrated that this issue needs to be integrated with other public health initiatives, for example, campaign-based and randomized testing of individuals (presence of pathogen or antibodies), clinical case reporting, and mobile-based contact-tracing, and self-reporting systems.

In Bangladesh, only Dhaka consists of a proper sewage facility which is prevailed by 20% population whereas, every other urban and rural areas depend on on-site facilities. Therefore, the sampling of the study was divided into two methods based on the availability of sewage facilities. For the urban areas that contain off-site sanitation facilities, the sampling locations were selected grid-based. Firstly, we identified the drainage system and with the help of GIS, we have divided the drainage system into grids to get a whole picture of the clustered houses or public places near the drain. In this way, we have collected samples from the drains of the Isolation Center, Hospitals, Prisons, Community Drain, City Drain, etc. However, in the case of rural settlements and places without a drainage system, the sampling procedure for selecting sites was different as they primarily depend on on-site sanitation facilities. Hence, to represent the whole community, we selected some nodal points which are public places such as mosques, markets, communal ponds, communal canals, rail stations, bus stations, etc. These points were selected based on representing the community of an area.

Based on RT-PCR analysis of the water samples, in more than 92% of cases, we found the presence of the SARS-COV-2 gene (ORF1ab, N, and Internal Control-IC). However, The Ct value for different sampling locations and first and second-round sampling were not found similar. The cumulative Ct values for three different genes (ORF1ab, N, and IC) were found lower in the first-round sampling in December 2020. However, the concentration of the same genes was found higher in the second round of sampling and correlated with higher positive laboratory-confirmed cases. Several previous studies also had the same findings(Romero-Alvarez et al., 2021; Zacharioudakis et al., 2020). Therefore, based on our research findings, Ct values can be used as a tool to determine the trends of viral load which ultimately will help in identifying the number of patients without starting the clinical diagnosis process.

When the first and second round viral load were compared, the spread of genetic material in the on-site sanitation system was found more prominent than that of off-site sewage. It indicated the frequent spread of genetic from urban to rural areas with no adequate sewage system. Therefore, the findings from analyzing Ct values can also serve as an early warning tool for areas not affected or having asymptomatic cases. However, this variable trend of Ct values demands the necessity of analyzing wastewater samples after a certain interval for a longer period. The influence of seasonal variation also needs to be considered as it plays an important role in the spread of pathogenic viruses/bacteria(Liu et al., 2021; Pramanik et al., 2020; Rayan, 2021). The dynamics of viral shedding in feces, viral persistence in the sewer network, variation in wastewater flow, pH, and temperature due to climatic conditions are some of the important factors which played a significant role in this regard. (Challener et al., 2020; Holmdahl and Buckee, 2020; Roda et al., 2020; Lamarre and Talbot, 1989; Mecenas et al., 2020; Sajadi et al., 2020; Zhao et al., 2020). Spatiotemporal monitoring allowed the trend monitoring of SARS-CoV-2 transmission which could be used as a tool to assess the public health status (Bertacchini et al., 2020; Ling et al.). The methodology can also be applied in places where the risk is very high such as airports, shopping malls, and water transport-related sewage monitoring.

Here the interesting fact is that the SARS-CoV-2 genetic material distribution and their spreading potentials are not the same for the entire country. The presence of SARS-COV-2 genetic material was found low in sewer water despite the high population density in the refugee camp. The genetic make-up of the SARS-CoV-2 variants themselves might have influences due to extended environmental stability than human genes (Azuma et al., 2020; Riddell et al., 2020). The lowest existence of IC gene and the highest occurrence of ORF1ab followed by N gene in both (1^st^ time and 2^nd^ time) sampling rounds indicates that genetic materials of SARS-COV-2 were more resistant to damage than that of the human gene in wastewater. However, to understand the issue better, further research is required in delineating implications of host genetics, lifestyle, environmental, and other socioeconomic factors. Besides, extended research on circulating SARS-CoV-2 strains can be carried out simultaneously for a better understanding of SARS-COV-2 pandemics in the refugee community.

Moreover, the determination of the prevalence of SARS-CoV-2 is highly dependent on the characteristics of a specific location, demographic characteristics, and also on physicochemical parameters (Akter et al., 2020; Bhattacharya et al., 2021; Nguyen et al., 2020; Pandey et al., 2021; Rahimi et al., 2021). Furthermore, an unplanned sewage system with frequent Covid-hazard exposure might create a complicated scenario in the developing world. As per the considerations of global health experts, living with a huge population including various slum areas apprehend massive spreading of the SARS-COV-2 genetic materials resulting in the highest asymptomatic and confirmed cases (Anwar et al., 2020; Sharifi and Khavarian-Garmsir, 2020)

In the present study, we considered all these confounders to elucidate the relationship between quantitative Ct values and the probable number of SARS-COV-2 patients in a specific community. A SARS-COV-2 specific dashboard could also be developed, where the Ct values can be illustrated in trend graphs for different regions. Ct values can also be represented by a quantitative value, to identify the number of gene copies which is also an internationally accepted method (e.g., poliovirus surveillance in wastewater). The main source of the genetic material of SARS-COV-2 in wastewater is accumulated from stool, urine, and cough or the body fluids of symptomatic and asymptomatic patients. Therefore, the study findings presented indicate that if the number of positive cases increases, the concentration of genetic materials in wastewater will also increase.

With the novel findings of the present two-steps sampling study, the challenging journey towards the development of a model to predict patient numbers using the viral titers from wastewater with on-site sanitation facilities in developing countries will be possible. It was also observed from our study that the total bacterial count was found highest in the wastewater of Dhaka city than that of the wastewater samples collected from other study areas. Further research to identify other parameters will facilitate the development of the WBE surveillance system in the country. Furthermore, the relationship between SARS-COV-2 confirmed cases and genetic materials from wastewater emphasizes the need for taking the strongest precaution in treating sewer water of isolation centers. Untreated sewage flow from isolation centers can create problems through horizontal gene transfer with other organisms of the ecosystem. However, further research is required to investigate the detrimental effect of persisting SARS-CoV-2 genetic materials in the wastewater.

Bangladesh is currently passing through the second stage of the SARS-COV-2 pandemic. However, the experience of other countries demonstrates that the second or even third waves of infection are possible (Fisayoand Tsukagoshi, 2021; Graichen, 2021) which might create serious consequences in managing community health. Considering the upcoming situation, the best approach would be to develop a wastewater-based surveillance system for the entire country.(Barceló, 2020; Bivins et al., 2020; Mao et al., 2020; Polo et al., 2020; Street et al., 2020)

## 5. Conclusion

We focused on on-site drainage water-based surveillance system and demonstrated the detection of SARS-CoV-2 genetic materials in the wastewater and are planning to establish the surveillance system as a model for developing countries such as Bangladesh. This is the first study to demonstrate that wastewater-based epidemiology may be used to detect the prevalence of SARS-COV-2 in Bangladesh utilizing the ORF1ab and N genes of SARS-CoV-2. It can be used as a prediction tool which was discovered when the first and second round viral load were compared and was found the spread of genetic material in on-site sanitation systems being more prominent than that of the off-site sewage system. It was discovered that the on-site sewage systems of isolation centers hold a large proportion of SARS-CoV-2 genetic components after comparing different wastewater locations in Bangladesh. The approach was sensitive enough to detect SARS-CoV-2 RNA in the sewer system in places where SARS-COV-2 cases had been documented, such as a SARS-COV-2 isolation center, hospital community drain, city drain, communal pond, river, and mosques. The association of pH and bacterial load with the confirmed cases and genetic material was identified. The results showed good resolution and gave a clear indication of SARS-COV-2 patient loadings’ temporal variation. Even though the case numbers correspond to higher RNA concentrations in wastewater, the temporal changes in SARS-CoV-2 RNA concentrations need to be examined further from the daily, short-term, and long-term perspectives. To ensure treatment availability and assess the severity of the infection, it is necessary to study high-prevalence locations. The proposed study will help to understand the severity of transmission through wastewater in areas consisting of both off-site and on-site sanitation systems.

## Data Availability

Data will be made available on request.

## References

Ahmed, F., Islam, M. A., Kumar, M., Hossain, M., Bhattacharya, P., Islam, M. T., Hossen, F., Hossain, M. S., Islam, M. S., Uddin, M. M., Islam, M. N., Bahadur, N. M., Didar-ul-Alam, M., Reza, H. M., and Jakariya, M. (2021). First detection of SARS-CoV-2 genetic material in the vicinity of COVID-19 isolation Centre in Bangladesh: Variation along the sewer network. Science of The Total Environment, 776, 145724. https://doi.org/10.1016/j.scitotenv.2021.145724

Ahmed, W., Angel, N., Edson, J., Bibby, K., Bivins, A., O’Brien, J. W., Choi, P. M., Kitajima, M., Simpson, S. L., Li, J., Tscharke, B., Verhagen, R., Smith, W. J. M., Zaugg, J., Dierens, L., Hugenholtz, P., Thomas, K. V., and Mueller, J. F. (2020). First confirmed detection of SARS-CoV-2 in untreated wastewater in Australia: A proof of concept for the wastewater surveillance of COVID-19 in the community. Science of the Total Environment, 728, 138764. https://doi.org/10.1016/j.scitotenv.2020.138764

Ahmed, W., Bivins, A., Bertsch, P. M., Bibby, K., Choi, P. M., Farkas, K., Gyawali, P., Hamilton, K. A., Haramoto, E., Kitajima, M., Simpson, S. L., Tandukar, S., Thomas, K. V., and Mueller, J. F. (2020). Surveillance of SARS-CoV-2 RNA in wastewater: Methods optimization and quality control are crucial for generating reliable public health information. Current Opinion in Environmental Science and Health, 17, 82–93. https://doi.org/10.1016/j.coesh.2020.09.003

Akter, S., Roy, P. C., Ferdaus, A., Ibnat, H., Alam, A. S. M. R. U., Nigar, S., Jahid, I. K., and Hossain, M. A. (2020). Prevalence and stability of SARS CoV-2 RNA on Bangladeshi banknotes. MedRxiv, 2020.11.26.20233627. https://doi.org/10.1101/2020.11.26.20233627

Ammar, A., Trabelsi, K., Brach, M., Chtourou, H., Boukhris, O., Masmoudi, L., Bouaziz, B., Bentlage, E., How, D., Ahmed, M., Mueller, P., Mueller, N., Hammouda, O., Paineiras-Domingos, L., Braakman-jansen, A., Wrede, C., Bastoni, S., Pernambuco, C., Mataruna, L., … Hoekelmann, A. (2021). Effects of home confinement on mental health and lifestyle behaviours during the COVID-19 outbreak: Insight from the ECLB-COVID19 multicenter study. Biology of Sport, 38(1), 9–21. https://doi.org/10.5114/biolsport.2020.96857

Anwar, S., Nasrullah, M., and Hosen, M. J. (2020). COVID-19 and Bangladesh: Challenges and How to Address Them. Frontiers in Public Health, 8. https://doi.org/10.3389/fpubh.2020.00154

Arora, S., Nag, A., Sethi, J., Rajvanshi, J., Saxena, S., Shrivastava, S. K., and Gupta, A. B. (2020). Sewage surveillance for the presence of SARS-CoV-2 genome as a useful wastewater based epidemiology (WBE) tracking tool in India. Water Science and Technology, 82(12), 2823–2836. https://doi.org/10.2166/wst.2020.540

Azuma, K., Yanagi, U., Kagi, N., Kim, H., Ogata, M., and Hayashi, M. (2020). Environmental factors involved in SARS-CoV-2 transmission: effect and role of indoor environmental quality in the strategy for COVID-19 infection control. Environmental Health and Preventive Medicine, 25(1), 66. https://doi.org/10.1186/s12199-020-00904-2

Barceló, D. (2020). Wastewater-Based Epidemiology to monitor COVID-19 outbreak: Present and future diagnostic methods to be in your radar. Case Studies in Chemical and Environmental Engineering, 2, 100042. https://doi.org/10.1016/j.cscee.2020.100042

Bertacchini, F., Bilotta, E., and Pantano, P. S. (2020). On the temporal spreading of the SARS-CoV-2. PLoS ONE, 15(10 October). https://doi.org/10.1371/JOURNAL.PONE.0240777

Bhattacharya, P., Kumar, M., Islam, M. T., Haque, R., Chakraborty, S., Ahmad, A., Niazi, N. K., Cetecioglu, Z., Nilsson, D., Ijumulana, J., van der Voorn, T., Jakariya, M., Hossain, M., Ahmed, F., Rahman, M., Akter, N., Johnston, D., and Ahmed, K. M. (2021). Prevalence of SARS-CoV-2 in Communities Through Wastewater Surveillance—a Potential Approach for Estimation of Disease Burden. Current Pollution Reports, 7(2), 160–166. https://doi.org/10.1007/s40726-021-00178-4

Bivins, A., North, D., Ahmad, A., Ahmed, W., Alm, E., Been, F., Bhattacharya, P., Bijlsma, L., Boehm, A. B., Brown, J., Buttiglieri, G., Calabro, V., Carducci, A., Castiglioni, S., Cetecioglu Gurol, Z., Chakraborty, S., Costa, F., Curcio, S., de los Reyes, F. L., … Bibby, K. (2020). Wastewater-Based Epidemiology: Global Collaborative to Maximize Contributions in the Fight Against COVID-19. Environmental Science and Technology, 54(13), 7754–7757. https://doi.org/10.1021/acs.est.0c02388

Challener, D. W., Dowdy, S. C., and O’Horo, J. C. (2020). Analytics and Prediction Modeling During the COVID-19 Pandemic. Mayo Clinic Proceedings, 95(9), S8–S10. https://doi.org/10.1016/j.mayocp.2020.05.040

Chen, S., Prettner, K., Kuhn, M., Geldsetzer, P., Wang, C., Bärnighausen, T., and Bloom, D. E. (2021). Climate and the spread of COVID-19. Scientific Reports, 11(1), 9042. https://doi.org/10.1038/s41598-021-87692-z

Fisayo, T., and Tsukagoshi, S. (2021). Three waves of the COVID-19 pandemic. Postgraduate Medical Journal, 97(1147), 332–332. https://doi.org/10.1136/postgradmedj-2020-138564

Graichen, H. (2021). What is the difference between the first and the second/third wave of Covid-19ã – German perspective. Journal of Orthopaedics, 24, A1–A3. https://doi.org/10.1016/j.jor.2021.01.011

Haramoto, E., Malla, B., Thakali, O., and Kitajima, M. (2020). First environmental surveillance for the presence of SARS-CoV-2 RNA in wastewater and river water in Japan. Science of the Total Environment, 737, 140405. https://doi.org/10.1016/j.scitotenv.2020.140405

Holmdahl, I., and Buckee, C. (2020). Wrong but Useful — What Covid-19 Epidemiologic Models Can and Cannot Tell Us. New England Journal of Medicine, 383(4), 303–305. https://doi.org/10.1056/nejmp2016822

Kaplin, A., Junker, C., Kumar, A., Ribeiro, M. A., Yu, E., Wang, M., Smith, T., Rai, S. N., and Bhatnagar, A. (2021). Evidence and magnitude of the effects of meteorological changes on SARS-CoV-2 transmission. PLoS ONE, 16(2 February), e0246167. https://doi.org/10.1371/journal.pone.0246167

Kumar, M., Patel, A. K., Shah, A. V., Raval, J., Rajpara, N., Joshi, M., and Joshi, C. G. (2020). First proof of the capability of wastewater surveillance for COVID-19 in India through detection of genetic material of SARS-CoV-2. Science of the Total Environment, 746, 141326. https://doi.org/10.1016/j.scitotenv.2020.141326

La Rosa, G., Bonadonna, L., Lucentini, L., Kenmoe, S., and Suffredini, E. (2020). Coronavirus in water environments: Occurrence, persistence and concentration methods - A scoping review. Water Research, 179, 115899. https://doi.org/10.1016/j.watres.2020.115899

Lamarre, A., and Talbot, P. J. (1989). Effect of pH and temperature on the infectivity of human coronavirus 229E. Canadian Journal of Microbiology, 35(10), 972–974. https://doi.org/10.1139/m89-160

Ling, J., Hickman, R., Li, J., Lu, X., Lindahl, J., Viruses, Å. L.-, and 2020, undefined. (n.d.). Spatio-temporal mutational profile appearances of Swedish SARS-CoV-2 during the early pandemic. Mdpi.Com. https://doi.org/10.3390/v12091026

Liu, X., Huang, J., Li, C., Zhao, Y., Wang, D., Huang, Z., and Yang, K. (2021). The role of seasonality in the spread of COVID-19 pandemic. Environmental Research, 195, 110874. https://doi.org/10.1016/j.envres.2021.110874

Lodder, W., and de Roda Husman, A. M. (2020). SARS-CoV-2 in wastewater: potential health risk, but also data source. The Lancet Gastroenterology and Hepatology, 5(6), 533–534. https://doi.org/10.1016/S2468-1253(20)30087-X

Mao, K., Zhang, H., and Yang, Z. (2020). An integrated biosensor system with mobile health and wastewater-based epidemiology (iBMW) for COVID-19 pandemic. Biosensors and Bioelectronics, 169, 112617. https://doi.org/10.1016/j.bios.2020.112617

Mecenas, P., da Rosa Moreira Bastos, R. T., Rosário Vallinoto, A. C., and Normando, D. (2020). Effects of temperature and humidity on the spread of COVID-19: A systematic review. PLoS ONE, 15(9 September), e0238339. https://doi.org/10.1371/journal.pone.0238339

Medema, G., Heijnen, L., Elsinga, G., Italiaander, R., and Brouwer, A. (2020). Presence of SARS-Coronavirus-2 RNA in Sewage and Correlation with Reported COVID-19 Prevalence in the Early Stage of the Epidemic in the Netherlands. Environmental Science and Technology Letters, 7(7), 511–516. https://doi.org/10.1021/acs.estlett.0c00357

Nemudryi, A., Nemudraia, A., Wiegand, T., Surya, K., Buyukyoruk, M., Cicha, C., Vanderwood, K. K., Wilkinson, R., and Wiedenheft, B. (2020). Temporal Detection and Phylogenetic Assessment of SARS-CoV-2 in Municipal Wastewater. Cell Reports Medicine, 1(6), 100098. https://doi.org/10.1016/j.xcrm.2020.100098

Nguyen, T. T., Pham, T. N., Van, T. D., Nguyen, T. T., Nguyen, D. T. N., Le, H. N. M., Eden, J.-S., Rockett, R. J., Nguyen, T. T. H., Vu, B. T. N., Tran, G. Van, Le, T. Van, Dwyer, D. E., and van Doorn, H. R. (2020). Genetic diversity of SARS-CoV-2 and clinical, epidemiological characteristics of COVID-19 patients in Hanoi, Vietnam. PLOS ONE, 15(11), e0242537. https://doi.org/10.1371/journal.pone.0242537

Orive, G., Lertxundi, U., and Barcelo, D. (2020). Early SARS-CoV-2 outbreak detection by sewage-based epidemiology. Science of the Total Environment, 732, 139298. https://doi.org/10.1016/j.scitotenv.2020.139298

Pandey, U., Yee, R., Shen, L., Judkins, A. R., Bootwalla, M., Ryutov, A., Maglinte, D. T., Ostrow, D., Precit, M., Biegel, J. A., Bender, J. M., Gai, X., and Dien Bard, J. (2021). High Prevalence of SARS-CoV-2 Genetic Variation and D614G Mutation in Pediatric Patients With COVID-19. Open Forum Infectious Diseases, 8(6). https://doi.org/10.1093/ofid/ofaa551

Polo, D., Quintela-Baluja, M., Corbishley, A., Jones, D. L., Singer, A. C., Graham, D. W., and Romalde, J. L. (2020). Making waves: Wastewater-based epidemiology for COVID-19 – approaches and challenges for surveillance and prediction. Water Research, 186, 116404. https://doi.org/10.1016/j.watres.2020.116404

Prado, T., Fumian, T. M., Mannarino, C. F., Resende, P. C., Motta, F. C., Eppinghaus, A. L. F., Chagas do Vale, V. H., Braz, R. M. S., de Andrade, J. da S. R., Maranhão, A. G., and Miagostovich, M. P. (2021). Wastewater-based epidemiology as a useful tool to track SARS-CoV-2 and support public health policies at municipal level in Brazil. Water Research, 191, 116810. https://doi.org/10.1016/j.watres.2021.116810

Pramanik, M., Chowdhury, K., Rana, M. J., Bisht, P., Pal, R., Szabo, S., Pal, I., Behera, B., Liang, Q., Padmadas, S. S., and Udmale, P. (2020). Climatic influence on the magnitude of COVID-19 outbreak: a stochastic model-based global analysis. International Journal of Environmental Health Research, 1–16. https://doi.org/10.1080/09603123.2020.1831446

Prevost, B., Lucas, F. S., Goncalves, A., Richard, F., Moulin, L., and Wurtzer, S. (2015). Large scale survey of enteric viruses in river and waste water underlines the health status of the local population. Environment International, 79, 42–50. https://doi.org/10.1016/j.envint.2015.03.004

Rahimi, A., Mirzazadeh, A., and Tavakolpour, S. (2021). Genetics and genomics of SARS-CoV-2: A review of the literature with the special focus on genetic diversity and SARS-CoV-2 genome detection. Genomics, 113(1), 1221–1232. https://doi.org/10.1016/j.ygeno.2020.09.059

Randazzo, W., Truchado, P., Cuevas-Ferrando, E., Simón, P., Allende, A., and Sánchez, G. (2020). SARS-CoV-2 RNA in wastewater anticipated COVID-19 occurrence in a low prevalence area. Water Research, 181, 115942. https://doi.org/10.1016/j.watres.2020.115942

Rayan, R. A. (2021). Seasonal variation and COVID-19 infection pattern: A gap from evidence to reality. Current Opinion in Environmental Science and Health, 20, 100238. https://doi.org/10.1016/j.coesh.2021.100238

Riddell, S., Goldie, S., Hill, A., Eagles, D., and Drew, T. W. (2020). The effect of temperature on persistence of SARS-CoV-2 on common surfaces. Virology Journal, 17(1), 145. https://doi.org/10.1186/s12985-020-01418-7

Rimoldi, S. G., Stefani, F., Gigantiello, A., Polesello, S., Comandatore, F., Mileto, D., Maresca, M., Longobardi, C., Mancon, A., Romeri, F., Pagani, C., Cappelli, F., Roscioli, C., Moja, L., Gismondo, M. R., and Salerno, F. (2020). Presence and infectivity of SARS-CoV-2 virus in wastewaters and rivers. Science of the Total Environment, 744, 140911. https://doi.org/10.1016/j.scitotenv.2020.140911

Roda, W. C., Varughese, M. B., Han, D., and Li, M. Y. (2020). Why is it difficult to accurately predict the COVID-19 epidemicã Infectious Disease Modelling, 5, 271–281. https://doi.org/10.1016/j.idm.2020.03.001

Romero-Alvarez, D., Garzon-Chavez, D., Espinosa, F., Ligña, E., Teran, E., Mora, F., Espin, E., Albán, C., Galarza, J. M., and Reyes, J. (2021). Cycle Threshold Values in the Context of Multiple RT-PCR Testing for SARS-CoV-2. Risk Management and Healthcare Policy, Volume 14, 1311–1317. https://doi.org/10.2147/RMHP.S282962

Sajadi, M. M., Sajadi, M. M., Habibzadeh, P., Vintzileos, A., Shokouhi, S., Miralles-Wilhelm, F., Miralles-Wilhelm, F., Amoroso, A., and Amoroso, A. (2020). Temperature, Humidity, and Latitude Analysis to Estimate Potential Spread and Seasonality of Coronavirus Disease 2019 (COVID-19). JAMA Network Open, 3(6), e2011834. https://doi.org/10.1001/jamanetworkopen.2020.11834

Sharifi, A., and Khavarian-Garmsir, A. R. (2020). The COVID-19 pandemic: Impacts on cities and major lessons for urban planning, design, and management. Science of the Total Environment, 749, 142391. https://doi.org/10.1016/j.scitotenv.2020.142391

Sherchan, S. P., Shahin, S., Ward, L. M., Tandukar, S., Aw, T. G., Schmitz, B., Ahmed, W., and Kitajima, M. (2020). First detection of SARS-CoV-2 RNA in wastewater in North America: A study in Louisiana, USA. Science of the Total Environment, 743, 140621. https://doi.org/10.1016/j.scitotenv.2020.140621

Street, R., Malema, S., Mahlangeni, N., and Mathee, A. (2020). Wastewater surveillance for Covid-19: An African perspective. Science of The Total Environment, 743, 140719. https://doi.org/10.1016/j.scitotenv.2020.140719

Tang, X., Wu, C., Li, X., Song, Y., Yao, X., Wu, X., Duan, Y., Zhang, H., Wang, Y., Qian, Z., Cui, J., and Lu, J. (2020). On the origin and continuing evolution of SARS-CoV-2. National Science Review, 7(6), 1012–1023. https://doi.org/10.1093/nsr/nwaa036

Wu, F., Zhang, J., Xiao, A., Gu, X., Lee, W. L., Armas, F., Kauffman, K., Hanage, W., Matus, M., Ghaeli, N., Endo, N., Duvallet, C., Poyet, M., Moniz, K., Washburne, A. D., Erickson, T. B., Chai, P. R., Thompson, J., and Alm, E. J. (2020). SARS-CoV-2 Titers in Wastewater Are Higher than Expected from Clinically Confirmed Cases. MSystems, 5(4). https://doi.org/10.1128/msystems.00614-20

Wurtzer, S., Marechal, V., Mouchel, J. M., Maday, Y., Teyssou, R., Richard, E., Almayrac, J. L., and Moulin, L. (2020). Evaluation of lockdown effect on SARS-CoV-2 dynamics through viral genome quantification in waste water, Greater Paris, France, 5 March to 23 April 2020. Eurosurveillance, 25(50). https://doi.org/10.2807/1560-7917.ES.2020.25.50.2000776

Zacharioudakis, I. M., Zervou, F. N., Prasad, P. J., Shao, Y., Basu, A., Inglima, K., Weisenberg, S. A., and Aguero-Rosenfeld, M. E. (2020). Association of SARS-CoV-2 genomic load trends with clinical status in COVID-19: A retrospective analysis from an academic hospital center in New York City. PLOS ONE, 15(11), e0242399. https://doi.org/10.1371/journal.pone.0242399

Zhao, L., Qi, Y., Luzzatto-Fegiz, P., Cui, Y., and Zhu, Y. (2020). COVID-19: Effects of Environmental Conditions on the Propagation of Respiratory Droplets. Nano Letters, 20(10), 7744–7750. https://doi.org/10.1021/acs.nanolett.0c03331

