## Supplementary Materials for "Wastewater based surveillance system to detect SARS-CoV-2 genetic material for countries with on-site sanitation facilities: an experience from Bangladesh" (from another version of this preprint: version 1, 2021-08-01)

**Supplementary Table-1**: Sampling Locations

| **Sample Number** | **District** | **Source** | **Latitude** | **Longitude** |
| --- | --- | --- | --- | --- |
| Sample-1 | Mymensing | Isolation Center Drain Effluent | 24.7437194 | 90.4111709 |
| Sample-2 |  | Sadar Hospital Drain Effluent | 24.7447129 | 90.4094664 |
| Sample-3 |  | Medical College Drain Wastage | 24.2329541 | 90.8322791 |
| Sample-4 |  | Community Drain Effluent | 24.2993697 | 91.5117848 |
| Sample-5 |  | City Drain Effluent | 24.762572 | 90.4508283 |
| Sample-6 |  | Bus Stand Drain Effluent | 24.7465384 | 90.4093229 |
| Sample-7 |  | Pond/River | 24.7468126 | 90.4172393 |
| Sample-8 | Sylhet | Isolation Center Drain Effluent | 24.9032097 | 91.8572579 |
| Sample-9 |  | Sadar Hospital Drain Effluent | 24.9005396 | 91.8520254 |
| Sample-10 |  | Medical College Drain Wastage | 24.9030955 | 91.8697104 |
| Sample-11 |  | Community Drain Effluent | 24.8820263 | 91.871447 |
| Sample-12 |  | City Drain Effluent | 24.8859395 | 91.8810497 |
| Sample-13 |  | Pond/River | 24.8994062 | 91.856101 |
| Sample-14 | Rajshahi | Isolation Center Drain Effluent | 25.328547 | 89.5430594 |
| Sample-15 |  | Sadar Hospital Drain Effluent | 22.8094006 | 89.5727876 |
| Sample-16 |  | Medical College Drain Wastage | 24.3704138 | 88.5899355 |
| Sample-17 |  | Community Drain Effluent | 24.3703097 | 88.5832622 |
| Sample-18 |  | City Drain Effluent | 22.8168106 | 89.5438576 |
| Sample-19 |  | Prison Drain Effluent | 22.8168106 | 89.5438576 |
| Sample-20 | Rangpur | Isolation Center Drain Effluent | 25.7345806 | 89.2616246 |
| Sample-21 |  | Sadar Hospital Drain Effluent | 25.7235396 | 89.2572299 |
| Sample-22 |  | Medical College Drain Wastage | 25.7355102 | 89.2600018 |
| Sample-23 |  | Community Drain Effluent | 25.7413608 | 89.2541844 |
| Sample-24 |  | City Drain Effluent | 25.7355102 | 89.2600018 |
| Sample-25 |  | Prison Drain Effluent | 25.7413608 | 89.2541844 |
| Sample-26 | Gaibandha | Isolation Center Drain Effluent | 22.8132005 | 89.561587 |
| Sample-27 |  | Sadar Hospital Drain Effluent | 22.823388 | 89.5522344 |
| Sample-28 |  | SKS Hospital | 22.8129739 | 89.5640421 |
| Sample-29 |  | Community Drain Effluent | 22.8170072 | 89.5677315 |
| Sample-30 |  | City Drain Effluent | 22.8129739 | 89.5640421 |
| Sample-31 |  | Bus Stand Drain Effluent | 22.8170072 | 89.5677315 |
| Sample-32 | Dhaka | Mohammadpur 1 | 22.6947725 | 90.3698629 |
| Sample-33 |  | Mohammadpur 2 | 22.6937856 | 90.364171 |
| Sample-34 |  | Mohammadpur 3 | 22.6866324 | 90.3626151 |
| Sample-35 |  | Mohammadpur 4 | 23.8294594 | 90.3744785 |
| Sample-36 |  | Mohammadpur 5 | 23.8038746 | 90.3618174 |
| Sample-37 |  | Mohammadpur 6 | 23.7994571 | 90.3587234 |
| Sample-38 |  | Mohammadpur 7 | 23.8038746 | 90.3618174 |
| Sample-39 |  | korail slum 2 | 23.8038746 | 90.3618174 |
| Sample-40 |  | korail slum 3 | 23.8038746 | 90.3618174 |
| Sample-41 |  | Kurmitola General Hospital | 23.8038746 | 90.3618174 |
| Sample-42 |  | Kurmitola General Hospital | 22.6937856 | 90.364171 |
| Sample-43 |  | Kurmitola General Hospital | 22.6937856 | 90.364171 |
| Sample-44 |  | Mugdha Medical College and Hospital | 22.6937856 | 90.364171 |
| Sample-45 |  | Mugdha Medical College and Hospital | 22.6937856 | 90.364171 |
| Sample-46 |  | Mugdha Medical College and Hospital | 22.6937856 | 90.364171 |
| Sample-47 |  | Kuwait Moitri Hospital | 23.8705311 | 90.4040088 |
| Sample-48 |  | Kuwait Moitri Hospital | 23.8705311 | 90.4040088 |
| Sample-49 |  | Kuwait Moitri Hospital | 23.8705311 | 90.4040088 |
| Sample-50 | Barisal | Isolation Center Drain Effluent | 22.823388 | 89.5522344 |
| Sample-51 |  | Sadar Hospital Drain Effluent | 22.8166126 | 89.5433873 |
| Sample-52 |  | Medical College Drain Wastage | 21.2140495 | 92.1383388 |
| Sample-53 |  | Community Drain Effluent | 21.2072313 | 92.1449269 |
| Sample-54 |  | City Drain Effluent | 21.2140495 | 92.1383388 |
| Sample-55 |  | Bus Stand Drain Effluent | 21.2072313 | 92.1449269 |
| Sample-56 | Chittagong | Isolation Center Drain Effluent | 22.3668585 | 91.7800097 |
| Sample-57 |  | Sadar Hospital Drain Effluent | 22.9345997 | 90.8299029 |
| Sample-58 |  | Medical College Drain Wastage | 22.934116 | 90.8325748 |
| Sample-59 |  | Community Drain Effluent | 22.3614896 | 91.7977401 |
| Sample-60 |  | City Drain Effluent | 22.3608166 | 91.7841252 |
| Sample-61 | Khulna | Isolation Center Drain Effluent | 22.813151 | 89.5615517 |
| Sample-62 |  | Sadar Hospital Drain Effluent | 24.3695491 | 88.5826917 |
| Sample-63 |  | Medical College Drain Wastage | 22.8170763 | 89.5679443 |
| Sample-64 |  | Community Drain Effluent | 24.3680047 | 88.584496 |
| Sample-65 |  | City Drain Effluent | 22.8094006 | 89.5727876 |
| Sample-66 | Cox’s Bazar | Isolation Center Drain Effluent | 22.8132005 | 89.561587 |
| Sample-67 |  | Sadar Hospital Drain Effluent | 22.823388 | 89.5522344 |
| Sample-68 |  | Medical College Drain Wastage | 22.8171145 | 89.5674751 |
| Sample-69 |  | Community Drain Effluent | 22.8166126 | 89.5433873 |
| Sample-70 |  | City Drain Effluent | 22.8129739 | 89.5640421 |
| Sample-71 | Cumilla | Isolation Center Drain Effluent | 21.47550376 | 90.05704923 |
| Sample-72 |  | Sadar Hospital Drain Effluent | 21.47550376 | 90.05704923 |
| Sample-73 |  | Medical College Drain Wastage | 22.57250371 | 22.57250371 |
| Sample-74 |  | Community Drain Effluent | 21.47550376 | 90.05704923 |
| Sample-75 |  | City Drain Effluent | 23.87650379 | 23.87650379 |
| Sample-76 | Kishorgonj | Isolation Center Drain Effluent | 24.5758133 | 90.7402032 |
| Sample-77 |  | Sadar Hospital Drain Effluent | 24.746997 | 90.4173974 |
| Sample-78 |  | Medical College Drain Wastage | 24.5762246 | 90.6953107 |
| Sample-79 |  | Community Drain Effluent | 24.7447129 | 90.4094664 |
| Sample-80 |  | City Drain Effluent | 24.2329541 | 90.8322791 |
| Sample-81 | Rohingya Camp | Networking Inlet(Pipenetwork) | 22.8132005 | 89.561587 |
| Sample-82 |  | Lagoon-1 Outlet | 22.823388 | 89.5522344 |
| Sample-83 |  | Lagoon-2 Outlet | 22.8171145 | 89.5674751 |
| Sample-84 |  | Sludge -1 Outlet | 22.8166126 | 89.5433873 |
| Sample-85 |  | Estonian chamber | 22.8129739 | 89.5640421 |
| Sample-86 |  | Upflow filter-1 | 22.8170072 | 89.5677315 |
| Sample-87 |  | Upflow filter-2 | 21.2144768 | 92.1378941 |
| Sample-88 |  | Siphon Store | 21.2770739 | 92.0914011 |
| Sample-89 |  | Trickling filter | 22.8132005 | 89.561587 |
| Sample-90 |  | Polishing Point | 22.823388 | 89.5522344 |
| Sample-91 |  | Inlet | 22.8171145 | 89.5674751 |
| Sample-92 |  | Lime treated discharge | 22.8166126 | 89.5433873 |
| Sample-93 |  | Wastewater from tubewell drain | 22.8129739 | 89.5640421 |
| Sample-94 |  | Bathing outlet | 22.8170072 | 89.5677315 |
| Sample-95 |  | Household waste | 21.2144768 | 92.1378941 |
| Sample-96 |  | Patient sample | 21.2770739 | 92.0914011 |
| Sample-97 |  | Primary Health outlet | 22.8132005 | 89.561587 |
| Sample-98 |  | Drinking water primary Health Care | 22.823388 | 89.5522344 |
| Sample-99 |  | The community used wastewater common sewer | 22.8171145 | 89.5674751 |
| Sample-100 |  | The outlet from all community use | 22.8166126 | 89.5433873 |
| Sample-101 |  | Community Tubewell discharge | 22.8129739 | 89.5640421 |
| Sample-102 |  | Community Drinking water | 22.8170072 | 89.5677315 |
| Sample-103 |  | Hospital (Ph.D.)Outlet | 21.2144768 | 92.1378941 |
| Sample-104 |  | Camp 4 outlet | 21.2770739 | 92.0914011 |
| Sample-105 |  | Camp 8 outlet | 22.8171145 | 89.5674751 |
| Sample-106 |  | 8 East Camp Community Drain Discharge | 22.8166126 | 89.5433873 |
| Sample-107 |  | Waste from a local shop | 22.8129739 | 89.5640421 |
| Sample-108 |  | Drain outlet | 22.8170072 | 89.5677315 |
| Sample-109 |  | Local Shop | 21.2144768 | 92.1378941 |
| Sample-110 |  | Local Shop | 21.2770739 | 92.0914011 |
| Sample-111 | Brahmonbaria | Isolation Center Drain Effluent | 24.0457314 | 91.2266613 |
| Sample-112 |  | Sadar Hospital Drain Effluent | 24.762572 | 90.4508283 |
| Sample-113 |  | Hospital Drain Wastage | 24.7465384 | 90.4093229 |
| Sample-114 |  | Community Drain Effluent | 24.7468126 | 90.4172393 |
| Sample-115 |  | City Drain Effluent | 24.7468126 | 90.4172393 |
| Sample-116 | Habigonj | Isolation Center Drain Effluent | 24.5712942 | 91.6027849 |
| Sample-117 |  | Sadar Hospital Drain Effluent | 24.2993697 | 91.5117848 |
| Sample-118 |  | Community Drain Effluent | 24.8994062 | 91.856101 |
| Sample-119 |  | City Drain Effluent | 24.8992816 | 91.8670604 |

**Supplementary Figure:**

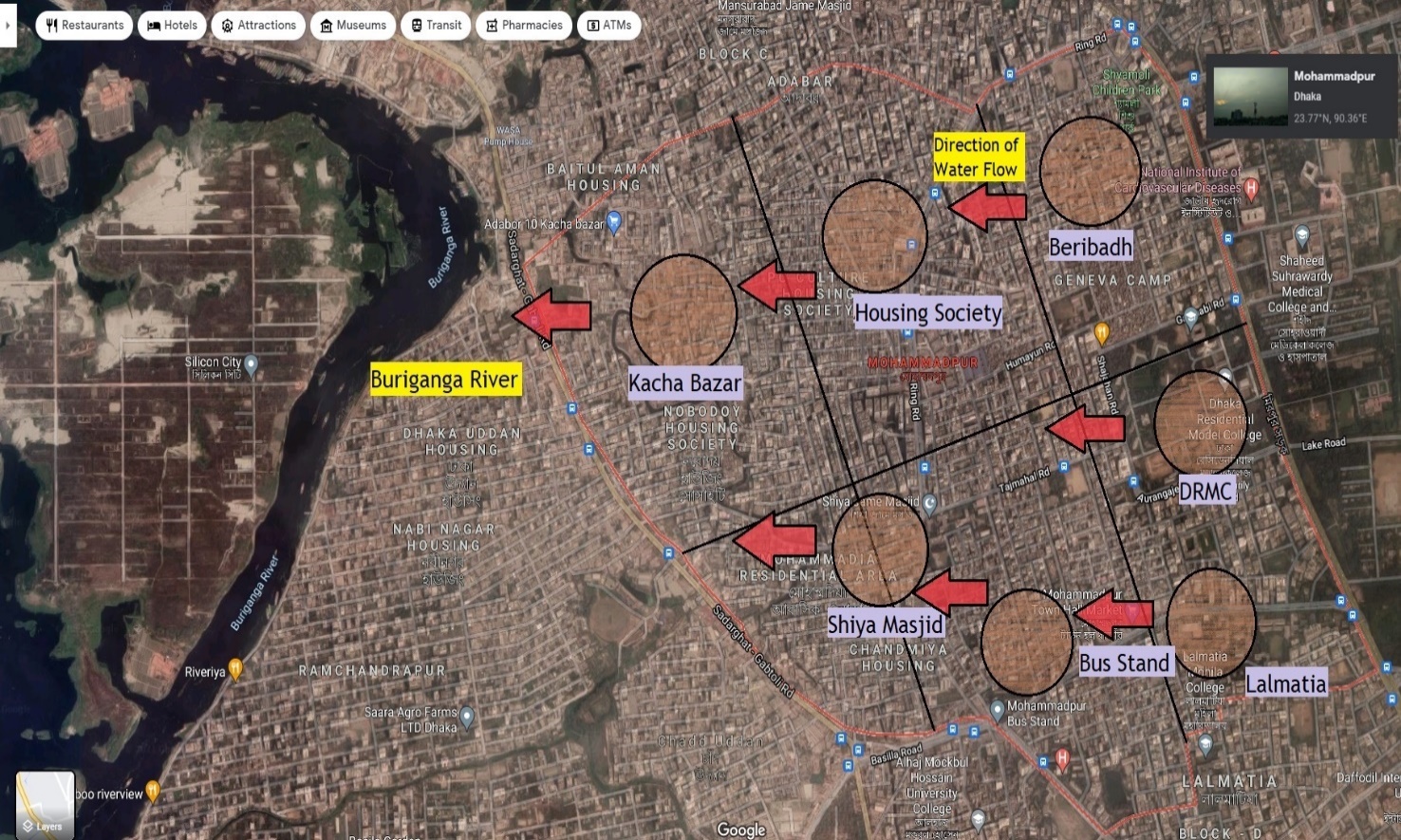
***Supplementary Figure-1****: Sample Collection from Mohammadpur, Dhaka, Bangladesh (on-site based epidemiology)*

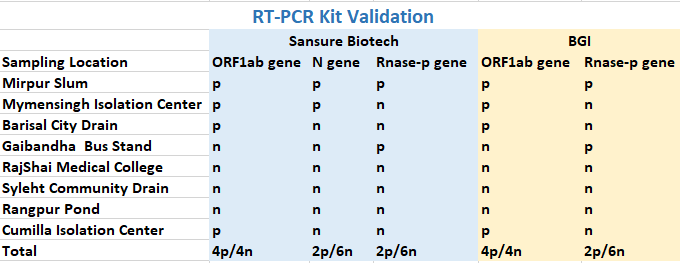

***Supplementary Figure-2:****RT-PCR result validation by another kit*
